# Comparisons of iron phenotypes and reports of menses, pregnancies, and live births in women with *HFE* p.C282Y homozygosity and *HFE* wt/wt

**DOI:** 10.64898/2026.01.02.25338237

**Authors:** James C. Barton, J. Clayborn Barton, Ronald T. Acton

## Abstract

**Introduction:** There is no comparison of iron phenotypes and menses, pregnancies, and live births reports of women with *HFE*-related hemochromatosis (*HFE* p.C282Y (rs1800562) homozygosity) and *HFE* wt/wt (absence of p.C282Y and *HFE* p.H63D (rs1799945)).

**Subjects and Methods:** We compared phenotypes and reports of non-Hispanic white women women aged ≥25 y in post-population screening evaluations using univariable methods.

**Results:** There were 153 p.C282Y/p.C282Y and 273 wt/wt. Median ages were 50 y (25, 86) and 55 y (25, 92), respectively (p=0.0019). Median transferrin saturation (TS), median serum ferritin (SF), and provisional iron overload prevalence were higher in p.C282Y/p.C282Y (p ≤0.0001, each comparison). Prevalences of documented iron overload (3.3% p.C282Y/p.C282Y vs. 0.7% wt/wt), iron overload-related disease (2.0% vs. 0.4%, respectively), and iron deficiency (3.9% vs. 2.6%, respectively) were not significantly different. Median ages at menarche (13 y p.C282Y/p.C282Y vs. 13 y wt/wt) and menopause (50 y vs. 49 y, respectively) were not significantly different. Reports of “in-between bleeding?” (24.2% p.C282Y/p.C282Y vs. 25.2% wt/wt, respectively), “early stopping of periods?” (11.8% vs. 13.9%, respectively), and “had a hysterectomy?” (30.1% vs. 35.9%, respectively) were not significantly different. Respective percentage pairs of women with p.C282Y/p.C282Y and wt/wt who reported 0, 1, 2, 3, or ≥4 pregnancies (or live births) did not differ significantly. Live births/pregnancies were 287/363 (79.1%, p.C282Y/p.C282Y) and 534/673 (79.3%, wt/wt) (p=0.7549).

**Conclusions:** Median TS, median SF, and provisional iron overload prevalence are greater in women with *HFE* p.C282Y/p.C282Y than those with wt/wt, although reports of menses, pregnancies, and live births are similar.

## Introduction

Hemochromatosis in persons of European descent is typically associated with homozygosity for p.C282Y (rs1800562), a common missense mutation of the homeostatic iron regulator (*HFE*, chromosome 6p22.2).^1,2^ HFE, a transmembrane glycoprotein, is an upstream regulator of the hepatic hormone hepcidin (*HAMP,* chromosome 19q13.12), the central regulator of iron homeostasis.^3^

The estimated prevalence of *HFE* p.C282Y homozygotes is 1 in 227 in non-Hispanic white persons in North America,^4^ 1 in 156 in persons of European descent in the United Kingdom,^5^ and 1 in 154 in persons of northern European descent in Australia.^6^ The percentages of women and men with p.C282Y homozygosity do not differ significantly.^4,6–9^ The combination of elevated transferrin saturation (TS), a surrogate marker of increased plasma iron transport, and elevated serum ferritin (SF), a surrogate marker of increased iron stores, was detected in 40-60% of untreated women with p.C282Y homozygosity who participated in population-based studies in the U.S.,^4,8^ Denmark,^9^ and Australia.^7^

Studies of women with *HFE* p.C282Y homozygosity have focused on the influence of menopause,^10,11^ numbers of pregnancies,^11,12^ and numbers of live births^11^ on iron phenotypes. There are few other descriptions of menses, pregnancies, and live births in women with p.C282Y homozygosity. Two girls with p.C282Y homozygosity aged 13 y and 16 y, respectively, had amenorrhea attributed to polycystic ovary syndrome (PCOS).^13^ A woman with p.C282Y homozygosity and mild von Willebrand disease had heavy menses.^14^ Another woman with p.C282Y homozygosity and elevated pre-conception TS and SF had a normal pregnancy and a healthy neonate.^15^ In a case-control study in the Netherlands, the prevalence of either p.C282Y homozygosity or p.C282Y heterozygosity in 157 women with pre-eclampsia and 157 control women did not differ significantly.^16^ In a population-based, mother-child cohort study in Norway, the risk of developing type 1 diabetes was not increased in the children of mothers with p.C282Y homozygosity or p.C282Y/p.H63D (rs1799945) compound heterozygosity.^17^

The aim of this study was to compare the aggregate iron phenotypes and questionnaire reports about menses, pregnancies, and live births of 153 self-identified non-Hispanic white women with *HFE* p.C282Y homozygosity with those of 273 self-identified non-Hispanic white women with *HFE* wt/wt (absence of p.C282Y and p.H63D) who participated in evaluations after primary care-based screening for hemochromatosis and iron overload.^18,19^ We discuss the present results in the context of previous studies of menses, pregnancies, and live births in women not selected for either hemochromatosis or p.C282Y homozygosity and in women diagnosed with hemochromatosis without documentation of *HFE*-related or non-*HFE*-related hemochromatosis genotypes.

## Methods

### Ethics approval statement

The Hemochromatosis and Iron Overload Screening (HEIRS) Study, conducted by the National Heart, Lung, and Blood Institute and the National Human Genome Research Institute, in accordance with the principles of the Declaration of Helsinki, evaluated diverse aspects of hemochromatosis, iron overload, and iron-related disorders in a primary care-based sample of 101,168 adults enrolled during the interval 2001-2003 at four Field Centers in the U.S. and one in Canada.^4^

Local Institutional Review Boards of the HEIRS Study Coordinating Center (Wake Forest University Institutional Review Board, Wake Forest University), the HEIRS Study Central Laboratory (University of Minnesota Institutional Review Board, University of Minnesota), and the HEIRS Study Field Centers (Medical Institutional Review Board, Howard University; UAB Institutional Review Board for Human Use, University of Alabama at Brimingham; University of California Irvine Institutional Review Board, University of California Irvine; Committee for the Protection of Human Subjects/Institutional Review Board, University of Oregon in collaboration with the University of Hawaii Biomedical Institutional Review Board, University of Hawaii/Honolulu; and London Health Sciences Centre Research Institute, London Health Sciences Centre) gave ethical approvals of the Study protocol that is described in detail elsewhere.^4,19,20^

### Participant consent statement

HEIRS Study participants ≥ 25 years of age were recruited from outpatient facilities affiliated with the five Field Centers and gave written informed consent for screening and post-screening evaluation.^19,20^ The HEIRS Study informed consent forms, not available as public documents, were used during the participant recruitment phase of the Study (2001-2002). Each of the five Field Centers in North America used an Institutional Review Board-approved consent form tailored to its specific institution.^19,20^

### Primary care-based screening

The HEIRS Study was designed as a cross-sectional, primary care-based screening study only.^4,19,20^ All participants reported race/ancestry categories approved by the National Heart, Blood, and Lung Institute and the National Human Genome Research Institute.^4,19,20^ Ninety-eight percent of self-identified non-Hispanic white participants were recruited at Field Centers in Alabama, California, Ontario, and Oregon/Hawaii.^4,19,20^ Laboratory testing included only TS and SF phenotyping and *HFE* p.C282Y and p.H63D allele-specific genotyping.^4,19,20^ *HFE* wt/wt was defined as the absence of both p.C282Y and p.H63D. Medical histories were not taken and physical examinations were not performed at the time of primary care-based screening.^4^

### Post-screening evaluations

Invitations to participate were extended to the following: 1) all *HFE* p.C282Y homozygotes; 2) all participants who had both elevated TS and elevated SF, regardless of *HFE* genotype; and 3) participants with wt/wt whose screening TS and SF were between the 25th and 75th percentiles of corresponding sex-specific distributions.^18^ Field Centers in the U.S. recruited 64.7% and 64.1% of women with p.C282Y homozygosity and *HFE* wt/wt, respectively, who attended post-screening evaluations (p = 0.9165).

Post-screening evaluations included the following: 1) questionnaires completed by participants that addressed medical histories, including questions about menses, pregnancies, and live births;^18,19,21^ 2) focused physical examinations performed by HEIRS Study physicians;^18,19^ and 3) laboratory testing of blood specimens.^18,19^ The median interval between screening and post-screening evaluations was eight months.^18^

### Selection of post-screening participants

We selected the 153 self-identified non-Hispanic white women with *HFE* p.C282Y homozygosity and the 273 self-identified non-Hispanic white women with wt/wt who fulfilled each of the following four criteria: 1) reported that they were not pregnant; 2) fasted overnight before attending evaluations; 3) answered medical history questions pertinent to the present study; and 4) had complete TS and SF data. We excluded the self-identified non-Hispanic white women who did not fulfill each of these four criteria (10 with p.C282Y homozygosity (6.1%) vs. 10 with wt/wt (3.5%), respectively; p = 0.2367).

### Post-screening laboratory testing

A morning blood sample was obtained after an overnight fast. Testing included TS, SF, serum alanine and aspartate aminotransferase (ALT, AST), and *HFE* genotyping (HEIRS Study Central Laboratory, Fairview-University Medical Center Clinical Laboratory, University of Minnesota, Fairview, MN, USA).^4^ The screening *HFE* genotypes were confirmed in each participant.

### Definitions of iron phenotypes

We adapted the iron overload phenotype classification of Allen et al.^6^ Provisional iron overload was defined as TS > 45% and SF > 200 µg/L.^6^ Documented iron overload was defined as SF > 1000 µg/L.^6^ Iron overload-related disease was defined as the combination of documented iron overload and at least one of the following: reported diagnosis of cirrhosis; reported diagnosis of liver cancer; swollen/tender 2nd/3rd metacarpophalangeal joints by physical examination; serum ALT > 40 IU/L; serum AST > 45 IU/L; and reported history of hemochromatosis.^6^ Obtaining liver biopsy specimens, making non-invasive estimates of hepatic iron content or fibrosis, performing quantitative phlebotomy, and evaluating participants for primary liver cancer were beyond the scope of the HEIRS Study. We defined iron deficiency as SF < 15 µg/L.^22^

### Statistics

The dataset for analyses consisted of observations on 426 women (153 *HFE* p.C282Y homozygotes, 273 *HFE* wt/wt). Data for age, TS, SF, numbers of pregnancies, and numbers of live births are displayed to the nearest integer. Kolmogorov-Smirnov testing demonstrated that these continuous variables differed significantly from those that are normally distributed. We displayed these data as medians (ranges) and compared them using Mann-Whitney U tests (two-tailed). We treated iron phenotype classifications and qualitative questionnaire reports as categorical data and compared them using the Fisher’s exact test (two-tailed) or the chi-square test (two-tailed), as appropriate. Percentages in some comparisons are displayed with 95% confidence intervals. We defined p < 0.05 to be significant. We used Excel^®^ 2021 (Microsoft Corp., Redmond, WA, USA) and GraphPad Prism 8^®^ (2018; GraphPad Software, San Diego, CA, USA).

## Results

### Characteristics of 426 women

There were 153 women with *HFE* p.C282Y homozygosity (35.9%) and 273 women with wt/wt (64.1%). The median age of women with p.C282Y homozygosity was lower than that of women with wt/wt (50 y (25, 86) vs. 55 y (25, 92), respectively; p = 0.0019). The respective percentages of women aged < 35 y did not differ significantly (11.1% p.C282Y homozygosity vs. 8.7% wt/wt; p = 0.4876).

### Iron phenotypes

Questionnaire reports indicated that 79.1% (121/153) of women with *HFE* p.C282Y homozygosity were not diagnosed with hemochromatosis before they participated in the HEIRS Study. Median TS, median SF, and prevalence of provisional iron overload were higher in women with p.C282Y homozygosity than those with wt/wt (Table 1).

**Table 1.**
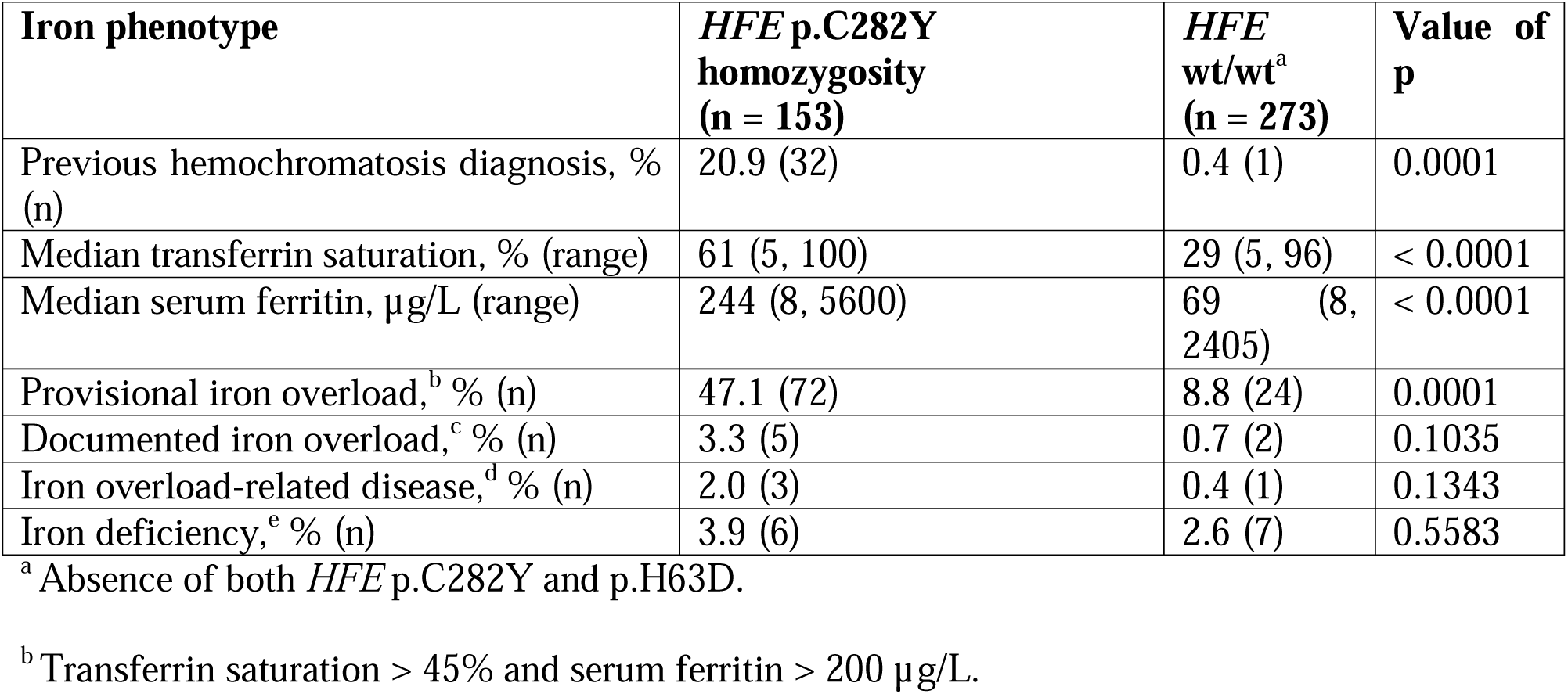
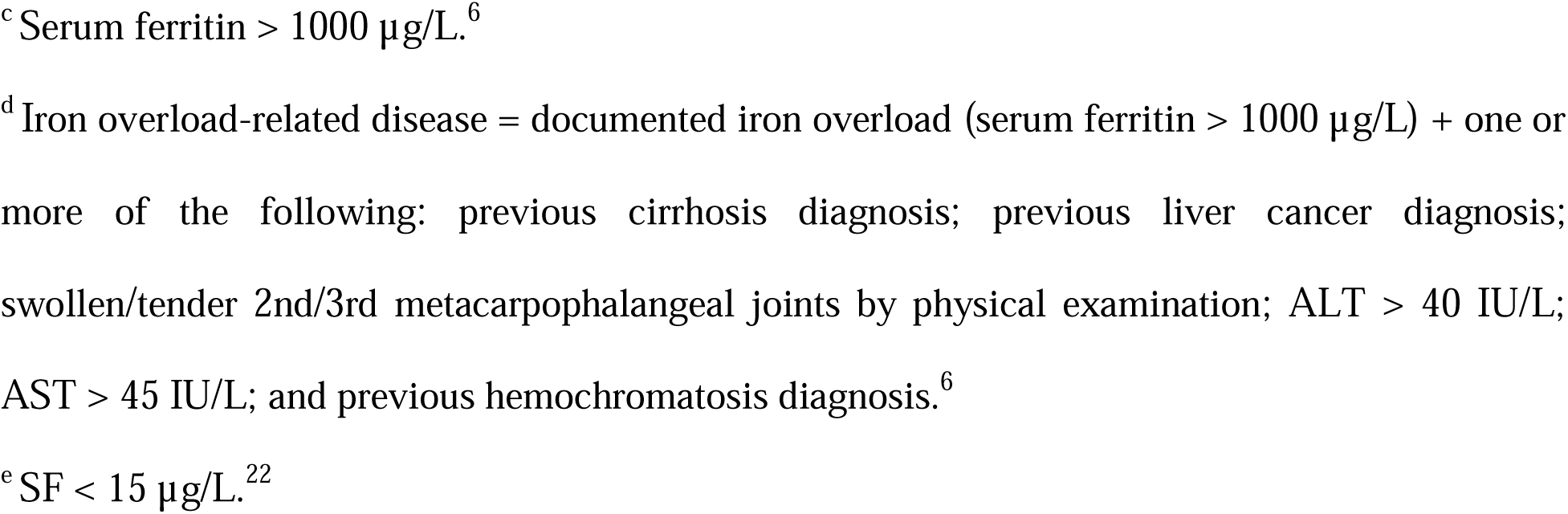
Iron phenotypes of 426 women in a post-screening study.

| Iron phenotype | *HFE* p.C282Y homozygosity (n = 153) | *HFE* wt/wt<sup>a</sup> (n = 273) | Value of p |
| --- | --- | --- | --- |
| Previous hemochromatosis diagnosis, % (n) | 20.9 (32) | 0.4 (1) | 0.0001 |
| Median transferrin saturation, % (range) | 61 (5, 100) | 29 (5, 96) | $< 0.0001$ |
| Median serum ferritin, $\mu\text{g/L}$ (range) | 244 (8, 5600) | 69 (8, 2405) | $< 0.0001$ |
| Provisional iron overload,<sup>b</sup> % (n) | 47.1 (72) | 8.8 (24) | 0.0001 |
| Documented iron overload,<sup>c</sup> % (n) | 3.3 (5) | 0.7 (2) | 0.1035 |
| Iron overload-related disease,<sup>d</sup> % (n) | 2.0 (3) | 0.4 (1) | 0.1343 |
| Iron deficiency,<sup>e</sup> % (n) | 3.9 (6) | 2.6 (7) | 0.5583 |

<sup>a</sup> Absence of both *HFE* p.C282Y and p.H63D.
<sup>b</sup> Transferrin saturation $> 45\%$ and serum ferritin $> 200 \mu\text{g/L}$ .
<sup>c</sup> Serum ferritin > 1000 µg/L.<sup>6</sup>
<sup>d</sup> Iron overload-related disease = documented iron overload (serum ferritin > 1000 µg/L) + one or more of the following: previous cirrhosis diagnosis; previous liver cancer diagnosis; swollen/tender 2nd/3rd metacarpophalangeal joints by physical examination; ALT > 40 IU/L; AST > 45 IU/L; and previous hemochromatosis diagnosis.<sup>6</sup>
<sup>e</sup> SF < 15 µg/L.<sup>22</sup>

Documented iron overload, iron overload-related disease, and iron deficiency were uncommon. The corresponding percentages of these iron phenotypes in women with *HFE* p.C282Y homozygosity and wt/wt did not differ significantly (Table 1).

### Menses

Median age at menarche, reports of abnormal menses, reports about hysterectomy, and median age at menopause in women with *HFE* p.C282Y homozygosity and wt/wt did not differ significantly (Table 2). The percentage of women with p.C282Y homozygosity who reported menopause was lower than that of women with wt/wt (Table 2). We attribute this difference to the lower median age of women with p.C282Y homozygosity.

**Table 2.** Menses questionnaire reports of 426 women in a post-screening study.

| **Report** | ***HFE* p.C282Y homozygosity (n = 153)** | ***HFE* wt/wt<sup>a</sup> (n = 273)** | **Value of p** |
| --- | --- | --- | --- |
| Median age at menarche, y (range) | 13 (9, 18) | 13 (8, 16) | 0.9045 |
| Yes to Menstrual problems?, % (n) | 45.8 (70) | 42.9 (117) | 0.6112 |
| Yes to In-between bleeding?, % (n) | 24.2 (37) | 25.6 (70) | 0.8160 |
| Yes to Early stopping of periods?, % (n) | 11.8 (18) | 13.9 (38) | 0.5542 |
| Yes to Had a hysterectomy?, % (n) | 30.1 (46) | 35.9 (98) | 0.2412 |
| Median age at hysterectomy, y (range) | 38 (22, 66) | 38 (21, 72) | 0.5157 |
| Yes to Gone through menopause?, % (n) | 52.3 (80/153) | 64.1 (175/273) | 0.0181 |
| Median age at menopause, y (range) | 50 (26, 59) (n = 80) | 49 (18, 71) (n = 175) | 0.7795 |
| Yes to Don't know about gone through menopause, % (n) | 6.5 (10) | 5.1 (14) | 0.6620 |

<sup>a</sup> Absence of both *HFE* p.C282Y and p.H63D.

Uncertainty about the occurrence of menopause was reported by 6.5% of women with *HFE* p.C282Y homozygosity and 5.1% of women with wt/wt (p = 0.6620) (Table 2). The median ages of these women were 50 y (41, 60) and 50 y (41, 69), respectively (p = 0.4839). We interpreted the combination of reports of uncertainty about menopause and participant ages as perimenopause.^23^

### Pregnancies

The percentages of women with *HFE* p.C282Y homozygosity and wt/wt who reported that they had “fertility problems” did not differ significantly (6.5% vs. 6.6%, respectively; p ∼1.0000). The respective percentage pairs of women with p.C282Y homozygosity or wt/wt who reported having 0, 1, 2, 3, or ≥ 4 pregnancies did not differ significantly (Figure 1).

**Figure 1.**
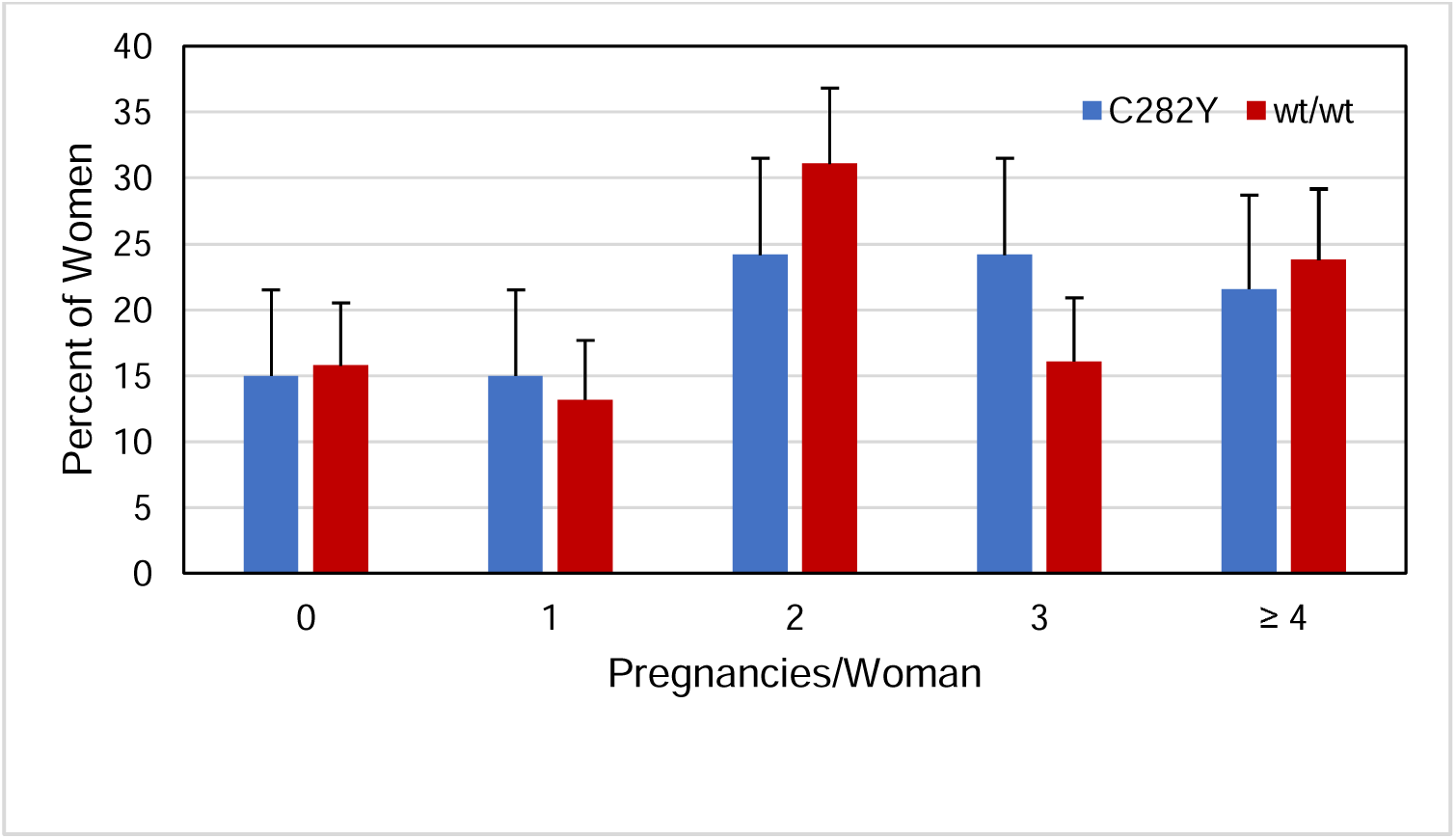
Pregnancies reported by 153 women with *HFE* p.C282Y homozygosity and 273 women with *HFE* wt/wt. These respective percentage pairs did not differ significantly (5 x 2 X^2^ = 5.4976; p = 0.2399). Error bars represent upper 95% confidence limits.

In 153 women with *HFE* p.C282Y homozygosity, the number of pregnancy reports was 363 (median 2 pregnancies/woman (0, 8)). In 273 women with wt/wt, the number of pregnancy reports was 673 (median 2 pregnancies/woman (0, 9)). The numbers of pregnancy reports by the numbers of women with p.C282Y homozygosity (363/153 = 2.3) and wt/wt (673/273 = 2.5) did not differ significantly (p = 0.7635).

### Live births

The respective percentage pairs of women with *HFE* p.C282Y homozygosity or wt/wt who reported having 0, 1, 2, 3, or ≥ 4 live births did not differ significantly (Figure 2). Two women with p.C282Y homozygosity and no women with wt/wt reported more live births than pregnancies (1.3% vs. 0%, respectively; p = 0.1392).

**Figure 2.**
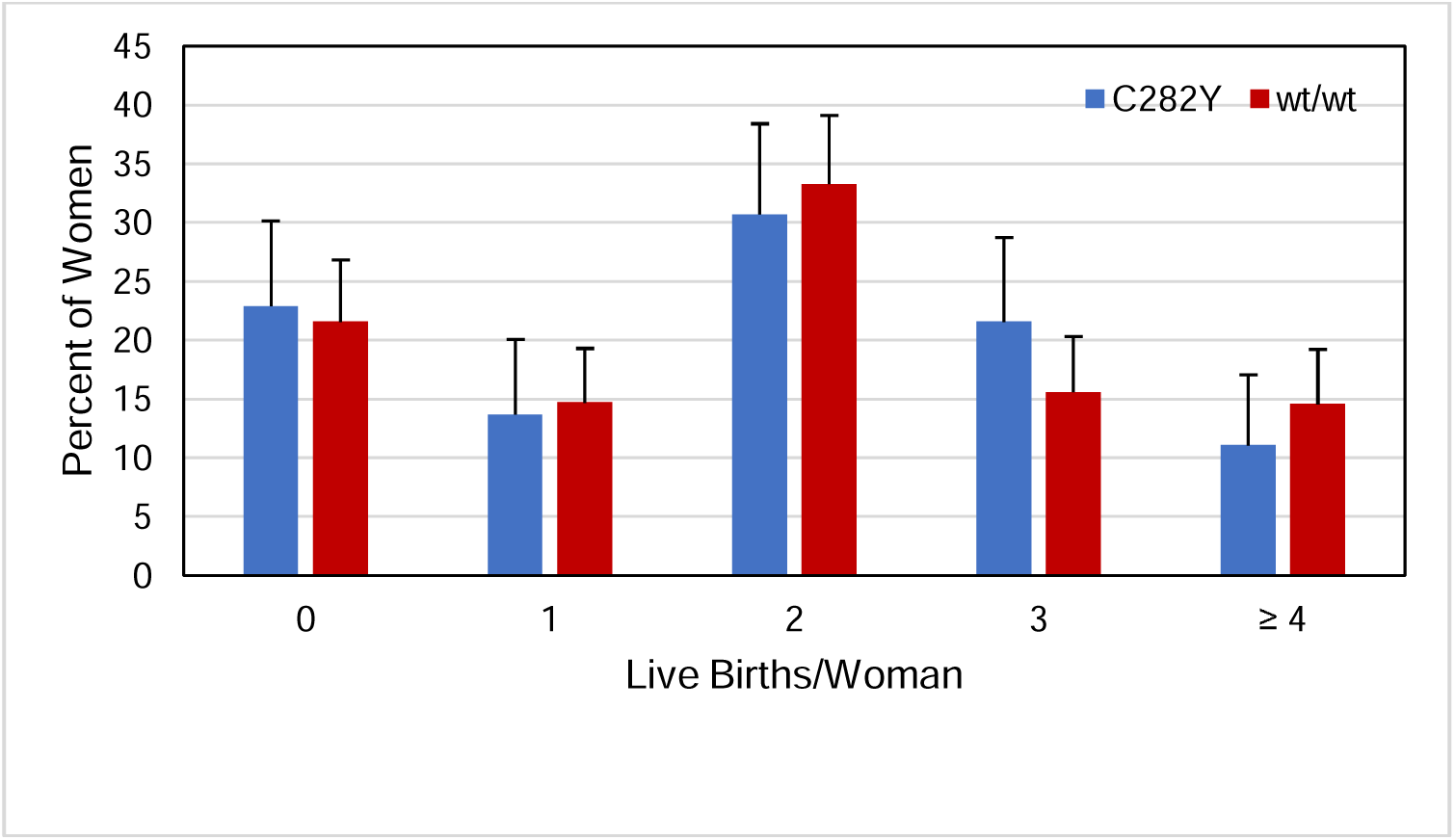
Live births reported by 153 women with *HFE* p.C282Y homozygosity and 273 women with *HFE* wt/wt. These respective percentage pairs did not differ significantly (5 x 2 X^2^ = 3.1156; p = 0.5387). Error bars represent upper 95% confidence limits.

In 153 women with *HFE* p.C282Y homozygosity, the number of live birth reports was 287 (median 2 live births/woman (0, 6)). Of reported pregnancies, 79.1% resulted in live birth reports. In 273 women with wt/wt, the number of live birth reports was 534 (median 2 live births/woman (0, 9)). Of reported pregnancies, 79.3% resulted in live birth reports. The numbers of live birth reports by the numbers of women with p.C282Y homozygosity (287/153 = 1.88) and wt/wt (534/273 = 1.96) did not differ significantly (p = 0.7549).

## Discussion

The strength and novelty of this study are the comparisons of aggregate iron phenotypes and aggregate questionnaire reports about menses and numbers of pregnancies and live births in 153 non-Hispanic women with *HFE* p.C282Y homozygosity and 273 non-Hispanic women with wt/wt. The absence of significant differences in aggregate questionnaire reports between these two cohorts of women indicates that the present reports were not significantly influenced by the *HFE* genotypes or the iron phenotypes we studied.

The prevalence of provisional iron overload in the present women with *HFE* p.C282Y homozygosity was greater than that of women with wt/wt, as expected,^6^ although the prevalences of documented iron overload, iron overload-related disease, and iron deficiency were low and did not differ significantly between the two subgroups of women. It is unknown whether the present women with wt/wt who also had documented iron overload and iron overload-related disease had non-*HFE*-related hemochromatosis or iron overload due to non-hemochromatosis disorders.

The median ages at menarche reported by the present women with *HFE* p.C282Y homozygosity and wt/wt agree with the median age at menarche in U.S. non-Hispanic white girls not selected for p.C282Y homozygosity.^24^ The percentages of hysterectomy reports and the median ages at hysterectomy of the present women with p.C282Y homozygosity and wt/wt are consistent with hysterectomy reports in non-Hispanic white women who participated in the 2021 National Health Interview Survey.^25^ The reports we interpreted as perimenopause^23^ are similar to those of other non-Hispanic white women in the U.S.^26^ The median ages of menopause in the present women with p.C282Y homozygosity and wt/wt were similar to those in a multi-ethnic sample of U.S. women who were not selected for *HFE* genotypes,^27^ although the HEIRS Study post-screening questionnaire did not permit distinguishing natural from surgical menopause.

The average numbers of live births per woman with *HFE* p.C282Y homozygosity and wt/wt in this study were similar to those in U.S. non-Hispanic white women who were not selected for *HFE* genotypes in 2000.^28^ The ratios of live births by pregnancies in this study did not differ between women with p.C282Y homozygosity and women with wt/wt. In U.S. non-Hispanic white women who were not selected for *HFE* genotypes in 2000, the ratio of live births by pregnancies was similar to that in the present women.^29^

Hemochromatosis was undiagnosed in 79.1% of the present women with *HFE* p.C282Y homozygosity before they participated in the HEIRS Study. Thus, gynecologists and obstetricians may not be aware that some non-Hispanic white women in their practices have hemochromatosis or p.C282Y homozygosity.

It was the consensus of a panel of experts chosen by the European Association for the Study of the Liver that iron deficiency should be treated before and during pregnancy in women with hemochromatosis^30^ because iron deficiency is a risk factor for adverse maternal and fetal outcomes.^31^ Advanced hepatic fibrosis and cirrhosis should be considered in pregnancy management,^30^ although these complications of iron overload due to *HFE* p.C282Y homozygosity in women are rare.^6^ There are no reports of prospective studies of pregnancy management or maternal and fetal outcomes in pregnant women with p.C282Y homozygosity.^30^

In a retrospective study of maternal and perinatal outcomes of 36 million delivery hospitalizations in the U.S. between 2010 and 2019, Niu et al. concluded that there were higher rates of “hypertensive disorders of pregnancy and venous thromboembolism among women with hemochromatosis, despite unaffected perinatal outcomes.”^32^ In a systematic review of published cases, Shamas concluded that maternal hemochromatosis was associated with “foetomaternal risks due to genetic predisposition.”^33^ Neither of these conclusions^32,33^ was based on studies of women who were selected because they had *HFE* p.C282Y homozygosity or other hemochromatosis-related genotypes.^3^

A University of Iowa case series study (2002-2006) revealed that only 153 of 601 hospitalized adult patients (25.5%) diagnosed with hemochromatosis had undergone *HFE* mutation analysis and were proven to be *HFE* p.C282Y homozygotes. Of the hospital patients misdiagnosed with hemochromatosis, 68% had known liver disease, and 5% had a hematologic cause of abnormal iron phenotypes.^34^ Together, these observations substantiate that misdiagnosis of hemochromatosis due to p.C282Y homozygosity in hospitalized adults is common.

Limitations of the present study include the lack of observations of women aged < 25 y or women other than self-identified non-Hispanic white women. HEIRS Study physicians did not interview post-screening participants about their past medical histories, review participant medical records, or perform gynecologic evaluations, by Study design. There were no post-screening evaluation questionnaire or physical examination items that provided information about the following: frequency of or quantities of blood lost from menses; assisted reproductive technology; complications of pregnancy and delivery; rates of abortion or fetal loss; health of neonates; history of breastfeeding; indications for and routes of hysterectomy; or diagnoses of endometriosis, PCOS, uterine fibroids, or sexually transmitted infections.

Our combined data suggest that it is unlikely that the differences in median ages of the present cohorts of women significantly affected their aggregate questionnaire reports except the occurrence of menopause. *HFE* wt/wt women whose screening TS and SF were in the first and fourth quartiles of sex-specific distributions were not invited to post-screening evaluations, although their inclusion may have increased the prevalences of proven iron overload, iron overload-related disease, and iron deficiency. It is unknown whether or not the aggregate iron phenotypes we observed at post-primary care-based evaluations were representative of the same women at pregnancy and childbirth.

## Conclusions

We conclude that median TS, median SF, and provisional iron overload prevalence of non-Hispanic white women with *HFE* p.C282Y homozygosity are greater than those of non-Hispanic women with wt/wt, although reports of menses, pregnancies, and live births are similar.

## Acknowledgments

The following individuals recruited participants and performed data collection (2001-2003) and received compensation for their work: Ronald T. Acton (Department of Microbiology, University of Alabama at Birmingham, Birmingham, Alabama); Paul C. Adams (Department of Medicine, London Health Sciences Centre, London, Ontario, Canada); James C. Barton (Department of Medicine, University of Alabama at Birmingham, Birmingham, Alabama and Southern Iron Disorders Center, Birmingham, Alabama); John H. Eckfeldt (Department of Laboratory Medicine and Pathology, University of Minnesota, Minneapolis, Minnesota); Victor R. Gordeuk (Division of Hematology and Oncology, Department of Medicine, University of Illinois at Chicago, Chicago, Illinois); Emily Harris (Epidemiology and Genomics Research Program, Division of Cancer Control and Population Sciences, National Cancer Institute, National Institutes of Health, Bethesda, Maryland); Helen Harrison (The Western-Fanshawe Collaborative BScN Program, Fanshawe College, London, Ontario, Canada); Christine E. McLaren (Department of Epidemiology, University of California, Irvine, California); and Gordon D. McLaren (Division of Hematology/Oncology, Department of Medicine, University of California, Irvine, California and Department of Veterans Affairs Long Beach Healthcare System, Long Beach, California).

## Data availability statement

Summary data supporting the conclusions of this study are displayed herein. The National Heart, Lung, and Blood Institute provides controlled access to individual participant data through the Biologic Specimen and Data Repository Information Coordinating Center (BioLINCC) (https://biolincc.nhlbi.nih.gov/studies/heirs/). Data access requires registration, evidence of local institutional review board approval or certification of exemption from institutional review board review, and completion of a data use agreement. The National Heart, Lung, and Blood Institute does not permit investigators to submit data directly to journals, related repositories, or other sources. Parties interested in obtaining the data analyzed in the present study are referred to BioLINCC.

